# Paternal body mass index and offspring DNA methylation: findings from the PACE consortium

**DOI:** 10.1101/2020.03.10.20020099

**Authors:** Gemma C Sharp, Rossella Alfano, Akram Ghantous, Jose Urquiza, Sheryl L Rifas-Shiman, Christian M Page, Jianping Jin, Silvia Fernández-Barrés, Gillian Santorelli, Gwen Tindula, Paul Yousefi, Leanne Kupers, Carlos Ruiz-Arenas, Vincent WV Jaddoe, Dawn DeMeo, Serena Fossati, John Wright, Karen Huen, Maja Popovic, Ellen A Nohr, George Davey Smith, Johanna Lepeule, Andrea Baccarelli, Maria C Magnus, Wenche Nystad, Maribel Casas, Emily Oken, Siri E Håberg, Marina Vafeiadi, Theano Roumeliotaki, Martine Vrijheid, Monica C Munthe-Kaas, Brenda Eskenazi, Luca Ronfani, Nina Holland, Leda Chatzi, Helle Margrete Meltzer, Zdenko Herceg, Michelle Plusquin, Mariona Bustamante, Marie-France Hivert, Deborah A Lawlor, Thorkild IA Sørensen, Stephanie J London, Janine F Felix, Caroline L Relton

## Abstract

**Background:** Accumulating evidence links paternal adiposity in the peri-conceptional period to offspring health outcomes. DNA methylation has been proposed as a mediating mechanism, but very few studies have explored this possibility in humans.

**Methods and findings:** In the Pregnancy And Childhood Epigenetics (PACE) consortium, we conducted a meta-analysis of co-ordinated epigenome-wide association studies (EWAS) of paternal prenatal Body Mass Index (BMI) (with and without adjustment for maternal BMI) in relation to DNA methylation in offspring blood at birth (13 datasets; total n= 4,894) and in childhood (six datasets; total n = 1,982). We found little evidence of association at either time point: for all CpGs, the False Discovery Rate-adjusted P-values were >0.05. In sex-stratified analyses, we found just four CpGs where there was robust evidence of association in female offspring. To compare our findings to those of other studies, we conducted a systematic review, which identified seven studies, including five candidate gene studies showing associations between paternal BMI/obesity and offspring or sperm DNA methylation at imprinted regions. However, in our own study, we found very little evidence of enrichment for imprinted genes.

**Conclusion:** Our findings do not support the hypothesis that paternal BMI around the time of pregnancy is associated with offspring blood DNA methylation, even at imprinted regions.

**Author Summary:** Previous small, mostly candidate gene studies have shown associations between paternal pre-pregnancy BMI and offspring blood DNA methylation. However, in our large meta-analysis of co-ordinated EWAS results from a total of 19 datasets across two timepoints, we found little evidence to support these findings, even at imprinted regions. This does not rule out the possibility of a paternal epigenetic effect in different tissues, at regions not covered by the 450k array, via different mechanisms, or in populations with greater extremes of paternal BMI. More research is warranted to help understand the size and nature of contributions of paternal adiposity to offspring epigenetics and health outcomes.

## Introduction

Accumulating evidence links paternal exposures in the peri-conceptional period to offspring health outcomes(1–3). Results from animal studies support a causal role for “paternal effects”(4) that are independent of maternal effects(2). Whereas prenatal maternal effects are most commonly postulated to occur via fetal intrauterine exposure, the suggested biological mechanisms underlying paternal effects(3,5) include germline de novo genetic mutations(6) or epigenetic changes(7), or alterations of components or properties of semen(8).

Epigenetic mechanisms that have been studied in relation to paternal exposures include DNA methylation, histone modification and microRNA expression(7), all of which can induce mitotically-heritable alterations in gene expression without changes to the DNA sequence. At most loci, patterns of DNA methylation are erased shortly after fertilization to create totipotent cells. However, some loci (most notably imprinted regions) can evade erasure, thus raising the possibility for intergenerational paternal epigenetic inheritance(9,10).

A large proportion of the research on paternal effects has explored dietary and metabolic exposures in relation to offspring metabolism and adiposity, sometimes including exploration of the potential mediating role of DNA methylation in sperm and offspring. Animal models of high fat diet-induced paternal obesity and diabetes have found associations with impaired offspring development(11) and offspring metabolic phenotypes and DNA methylation in pancreatic islets(12). In humans, a recent systematic review(13) found conflicting evidence on the association between paternal body mass index (BMI) and offspring birthweight, and some evidence of an association with greater offspring BMI, weight or body fat mass in childhood. A small number of studies have also found links between paternal BMI and sperm or offspring neonatal blood DNA methylation, but these have been based on candidate genes and/or had very limited sample sizes(14–20).

In the Pregnancy And Childhood Epigenetics (PACE) consortium(21), we conducted meta-analyses of coordinated epigenome-wide association studies (EWAS) exploring prenatal paternal BMI in relation to genome-wide DNA methylation at birth (cord blood; 13 independent datasets across ten cohorts) and in childhood (peripheral blood; six datasets across nine cohorts). A major challenge in studies of paternal effects is the correlation between paternal and maternal phenotypes and exposures, which could be due to shared environments and/or assortative mating(22). Several studies (for example, 22–24), including a large PACE consortium study(26), have shown that maternal BMI is associated with variation in offspring DNA methylation. To help disentangle any paternal effect from a maternal effect, we adjusted paternal associations for maternal BMI. We also conducted additional analyses with maternal BMI as the main exposure (unadjusted and adjusted for paternal BMI) and compared results to our primary analyses where paternal BMI was the main exposure. Finally, we systematically reviewed the literature on associations between paternal BMI and offspring or sperm DNA methylation in humans and assessed whether our results were enriched for imprinted loci and other regions identified by previous studies.

## Methods

### Meta-analysis of epigenome-wide association studies (EWAS)

#### Participating cohorts

The EWAS meta-analysis at birth included data from 13 independent datasets from ten cohorts in PACE (n=4,894): The Avon Longitudinal Study of Parents and Children (ALSPAC)(27–29); two independent datasets from Born in Bradford (BiB)(30); Center for the Health Assessment of Mothers and Children of Salinas (CHAMACOS)(31); Generation R Study (GENR)(32,33); the Genetics of Overweight Young Adults(34) (GOYA; nested within the Danish National Birth Cohort); INfancia y Medio Ambiente (INMA)(35); three independent datasets from the Norwegian Mother and Child Cohort Study (MoBa1, MoBa2, MoBa3)(36,37); Project Viva (Viva)(38); and two independent datasets generated as part of the EXPOsOMICS(39) project Piccolipiù(40) and RHEA(41).

The EWAS meta-analysis at childhood included data from six datasets from nine PACE cohorts (n=1,982): ALSPAC; CHAMACOS; Generation R; INMA, Project Viva and Human Early Life Exposome study (HELIX)(42). HELIX is a sample containing childhood methylation data pooled from several other cohorts (BIB; EDEN(43); INMA; MoBa; and RHEA). We conducted a sensitivity analysis excluding HELIX, because of concerns about potential sample overlap (albeit at a different time-point) with the INMA sample.

Cohorts are summarised in Table 1 and more detailed methods for each cohort are provided in Supporting Information File S1.

**Table 1:** A summary of key information for participating cohorts.

| Cohort | Country and ancestry | Methylation time point and array |  | Paternal BMI data source |  |  | Maternal BMI data source |  |
| --- | --- | --- | --- | --- | --- | --- | --- | --- |
|  |  | Birth | Childhood | Measured | Self-reported* | Reported by mother* | Measured* | Self-reported* |
| **ALSPAC** | United Kingdom; Northern European | 450k | 450k | × | ✓ (first trimester) | × | × | ✓ (pre-pregnancy) |
| **BIB (Asian)** | United Kingdom; Pakistani | EPIC | × | × | ✓ (first trimester) | × | ✓ (first trimester) |  |
| **BIB (European)** | United Kingdom; Northern European | EPIC | × | × | ✓ (first trimester) | × | ✓ (first trimester) |  |
| **CHAMACOS** | United States; Mexican American | 450k | 450k | ✓ (around delivery) | × | × | Height only (first trimester) | Weight only (pre-pregnancy) |
| **Generation R** | The Netherlands; Northern European | 450k | 450k | ✓ (first trimester) | × | × | × | ✓ (pre-pregnancy) |
| **GOYA** | Denmark; Northern European | 450k | × | × | × | ✓ (18 months after delivery) | × | ✓ (pre-pregnancy) |
| **HELIX (BIB, EDEN, INMA, MoBa, Rhea)** | Mixed (UK, France, Spain, Lithuania, Norway, Greece); European | × | 450k | × | BIB, MoBa (first trimester), EDEN (third trimester) | INMA, Rhea (first trimester) | BIB (BMI, first trimester) INMA, Rhea (height only, first trimester), EDEN (height only, second trimester) | EDEN (weight only, pre-pregnancy), INMA, Rhea (weight only, first trimester), MoBa (BMI, first trimester) |
| **INMA** | Spain; Southern European | 450k | 450k | × | × | ✓ (first trimester) | Height (first trimester) | Weight (first trimester) |
| **MoBa 1** | Norway; Northern European | 450k | × | × | × | ✓ (first trimester) | × | ✓ (first trimester) |
| **MoBa 2** | Norway; Northern European | 450k | × | × | × | ✓ (first trimester) | × | ✓ (first trimester) |
| **MoBa 3** | Norway; Northern European | 450k | × | × | × | ✓ (first trimester) | × | ✓ (first trimester) |
| **Piccolipiù** | Italy; Southern European | 450k | × | × | × | ✓ (around delivery) | × | ✓ (around delivery) |
| **Project Viva** | United States; European | 450k | 450k | × | × | ✓ (first trimester) | × | ✓ (pre-pregnancy) |
| **Rhea** | Greece (Crete); Southern European | 450k | × | × | × | ✓ (first trimester) | Height (first trimester) | Weight (first trimester) |

\*timepoints (i.e. pre-pregnancy, first trimester, etc) refer to the period participants were asked to report on rather than when the questionnaire was completed

### Measurement of paternal and maternal BMI

Paternal and maternal BMI was calculated using either self-reported or measured height and weight of participants (Table 1). For paternal BMI, cohorts selected a timepoint as close to the time of pregnancy as possible. Where possible, inclusion was restricted to biological fathers, but paternity status was not ascertainable in all cohorts. For maternal BMI, cohorts used self-reported pre-pregnancy BMI or BMI measured in the early stages of pregnancy. BMI was calculated in kg/m^2^ and then standardised for each cohort by converting to internal Z-scores.

### Measurement of DNA methylation

Biological samples were either cord blood samples from neonates or peripheral blood samples from children. DNA methylation was measured using either the llumina Infinium® HumanMethylation450 (486,425 probes) or EPIC (866,553 probes) BeadChip assay (Table 1). Probes that were common to both arrays (maximum 453,008) were included in the meta-analysis. Each cohort conducted its own laboratory methods, quality control and normalisation, as detailed in Supporting Information File S1. All cohorts used normalised, untransformed methylation beta values on a scale of 0 to 1.

### Other covariates

Questionnaire data was used to derive the following covariates, which were included in all adjusted models regardless of whether the main exposure was paternal or maternal BMI: maternal and paternal age (years); maternal smoking status during pregnancy (preferred definition: smoking throughout pregnancy/no smoking in pregnancy or quitting in the first trimester, but some cohorts used any/no smoking in pregnancy); paternal smoking status during or prior to pregnancy (any smoking in this time/no smoking in this time); maternal parity (one or more previous pregnancies/no previous pregnancies); and paternal socioeconomic position (higher/lower). For the latter variable, precise definitions were cohort-specific but most cohorts used paternal educational attainment (if data on paternal socioeconomic position were not available, maternal socioeconomic position was used). In addition, some models were stratified by sex of the child. Cohort specific information on covariate definitions is provided in Supporting Information File S1.

Systematic differences between samples (for example, those influenced by technical batch) were addressed by generating 20 surrogate variables using the SVA(44,45) R package. The number of surrogate variables (i.e. 20) was estimated using the ALSPAC dataset, and then each cohort estimated 20 SVs using their own data. Cellular heterogeneity was addressed by estimating cell type proportions using the Houseman algorithm(46) and either a cord blood reference panel(47) or a peripheral blood reference panel(48), depending on the methylation time-point.

### Cohort-specific EWAS

Each cohort performed independent EWAS according to a common, pre-specified analysis plan and R script (available on our Open Science Framework site at doi:10.17605/OSF.IO/EBTW7). If cohorts had data for both timepoints, EWAS were performed separately for birth and childhood. Potential methylation outliers, thought to be introduced by a technical error or a rare SNP were identified and removed using the Tukey method as previously described(49).

Linear regression models, modelling offspring methylation as the outcome and parental BMI as the exposure, were applied to each CpG using the Limma R package(50). Two main models were run for both paternal and maternal BMI: 1) a basic model where paternal or maternal BMI associations were adjusted for estimated cell proportions, surrogate variables for technical batch, maternal and paternal age, maternal smoking status during pregnancy, paternal smoking status during or prior to pregnancy, paternal socioeconomic position and maternal parity; and 2) a model with additional adjustment for the other parent’s BMI. In secondary analyses, the mutually adjusted models (i.e. model 2) were run stratified by sex of the offspring. This is because, although sex is not a true confounder (i.e. it does not influence pre-pregnancy parental BMI), it is a major source of variation in methylation(51) and there is some literature to support paternal effects occurring in a sex-specific manner (52,53). All probes were annotated to the human reference genome version 19, build 37.

### Other cohort-specific analyses

To allow us to explore the extent to which paternal BMI is associated with offspring blood cell proportions (which is an important source of variation in methylation data, but also an interesting phenotype to study in its own right (54)), cohorts provided results (effect estimate, standard error, p-value) for linear regressions of each estimated cell type on paternal BMI. These were then meta-analysed using the R package metafor (55).

Cohorts also provided the Spearman coefficient and P-value for the correlation between maternal and paternal BMI to allow us to assess the likelihood of assortative mating as an explanation for our results.

### Meta-analysis

Fixed-effects meta-analysis weighted by the inverse of the variance was performed at the University of Bristol using METAL(56). A shadow meta-analysis was also conducted independently by an author at the University of Hasselt (Rossella Alfano). All code used to perform these analyses is provided on our Open Science Framework site at doi:10.17605/OSF.IO/EBTW7.

### The EWAS meta-analysis pipeline was as follows

1. Filter probes from cohort results files to remove probes that are not common to both the EPIC and 450k array, control and QC probes, probes on SNPs, cross-hybridizing probes according to Chen et al.(57), and probes on the sex chromosomes;
2. Perform quality checks of cohort results by plotting correlation matrices of effect estimates generated by different models, generating QQ plots and calculating Lambda values, plotting the distribution of effect estimates, and producing “precision plots” of 1/median standard error against the square root of the sample size for each cohort and model;
3. Conduct a fixed effects meta-analysis using METAL for each model;
4. Adjust meta-analysis P-values for multiple testing using the false discovery rate (FDR) method. The threshold used to define statistical evidence of association was FDR-adjusted P-value<0.05.
5. Perform checks of meta-analysed results by plotting a correlation matrix of effect estimates generated by different models, generating QQ plots and calculating Lambda values;
6. Conduct a leave-one-out analysis using the R package metafor(55) at sites with the smallest P-values. The leave-one-out survival criteria we specified are: when any single cohort is omitted, the meta-analysis effect estimate should be in the same direction, not attenuate substantially (arbitrarily defined as >20% change-in-estimate), and not have a confidence interval that crosses the null;
7. Conduct a meta-regression to explore the impact of average age at DNA sample collection on the childhood EWAS meta-analysis results;
8. Conduct a sensitivity analysis at the birth time point, excluding cohorts that collected information on paternal BMI based on maternal-report (GOYA, INMA, MoBa, Piccolipiu, Project Viva, Rhea), because of concern about measurement error.
9. Conduct a sensitivity analysis at the childhood time point excluding HELIX because of concerns about overlap between some individuals in this dataset and individuals in INMA.

### Comparison of results for maternal and paternal BMI

To assess whether paternal and maternal BMI are associated with offspring methylation to similar extents and with similar distributions throughout the genome, we compared effect estimates for the EWAS meta-analyses with and without mutual adjustment for the other parent’s BMI. At the top ten CpGs associated with paternal BMI with the smallest P-values, we calculated the Cochrane Q statistic to explore statistical evidence for differences between the maternal and paternal effect estimates.

To explore the extent to which maternal BMI might explain associations between paternal BMI and offspring methylation, we conducted Kolmogorov-Smirnov tests to assess enrichment of our EWAS meta-analysis results for CpGs previously found to be associated with maternal BMI in the PACE consortium (26).

### Systematic literature review

To identify previous human studies of paternal adiposity and offspring or sperm methylation, we performed a systematic search of PubMed using the R package RISmed(58). Search terms (Supporting Information File S2) were formed using intersections of terms related to paternity, methylation and adiposity. Duplicate PubMed IDs and ineligible article types (non-journal articles and reviews) were excluded. Titles/abstracts were manually screened to assess inclusion based on whether the study investigated paternal adiposity and germ cell/offspring methylation, and whether it did so in humans. Information on study design, exposure, outcome, tissue, sample size, species and key relevant findings was manually extracted from the full text of included articles.

### Testing for enrichment of candidate loci identified through the literature

To explore enrichment of our EWAS meta-analysis results for loci identified through our literature search, we tested whether the distribution of EWAS meta-analysis P-values at these regions deviates from a null (uniform) distribution using Kolmogorov-Smirnov tests, and compared the direction of effect estimate to what has been reported previously. To identify CpG probes falling within a particular gene, we defined the location of the gene according to the GeneCards database (https://www.genecards.org) and human genome version 19 build 37.

### Availability of data and code

All code used to generate our results is available on our Open Science Framework site at doi:10.17605/OSF.IO/EBTW7. We are not able to publicly share individual level data from participating cohorts due to issues with consent and ethics, however all summary statistics generated by meta-analysis are also available at doi:10.17605/OSF.IO/EBTW7.

## Results

### EWAS meta-analysis at birth

#### Cohort summaries

Thirteen independent datasets were included in the EWAS meta-analyses at birth. Table 2 summarises key characteristics of these cohorts (more details in Supporting Information File S3). Around 48% of the babies were female. In all cohorts, paternal BMI had a higher mean and a lower standard deviation compared to maternal BMI. There was moderate correlation between both parents’ BMIs (r=0.2) which might be explained by assortative mating and/or shared environment.

**Table 2.** A summary of sex of the child and parental BMI for each cohort in the birth meta-analysis.

| Study | N (N female, N male) | Mean paternal BMI in kg/m<sup>2</sup> (SD) | Percentage fathers with BMI≥30 (i.e. obese) | Mean maternal BMI in kg/m<sup>2</sup> (SD) | Percentage mothers with BMI≥30 (i.e. obese) | Spearman's correlation between paternal and maternal BMI (P-value) |
| --- | --- | --- | --- | --- | --- | --- |
| ALSPAC | 531 (267, 264) | 25.0 (3.0) | 6% | 22.6 (3.3) | 4% | 0.2 (7.6x10<sup>-7</sup>) |
| Born in Bradford (British Asian) | 70 (27*, 43) | 26.6 (5.3) | 17% | 26.2 (5.8) | 21% | 0.2 (1.1x10<sup>-1</sup>) |
| Born in Bradford (White British) | 115 (59, 56) | 27.5 (4.7) | 24% | 26.7 (6.1) | 24% | 0.4 (9.1x10<sup>-4</sup>) |
| CHAMACOS | 158 (80, 78) | 28.0 (4.2) | 32% | 26.5 (4.5) | 19% | 0.1 (1.1x10<sup>-1</sup>) |
| Generation R | 947 (476, 471) | 25.2 (3.2) | 7% | 23.2 (3.9) | 6% | 0.2 (8.8x10<sup>-8</sup>) |
| GOYA | 390 (190, 200) | 25.1 (3.1) | 6% | 23.4 (3.7) | 6% | 0.1 (7.1x10<sup>-3</sup>) |
| INMA | 352 (173, 179) | 25.8 (3.5) | 13% | 23.8 (4.5) | 9% | 0.2 (4.7x10<sup>-5</sup>) |
| MoBa1 | 982 (458, 524) | 25.6 (3.1) | 8% | 24.0 (4.1) | 9% | 0.1 (6.2*10<sup>-5</sup>) |
| MoBa2 | 621 (275, 346) | 25.7 (3.1) | 8% | 24.3 (4.6) | 11% | 0.2 (2.2*10<sup>-8</sup>) |
| MoBa3 | 212 (109, 103) | 26.0 (3.1) | 9% | 24.0 (3.9) | 9% | 0.3 (2.0*10<sup>-4</sup>) |
| Piccolipiu | 98 (45, 53) | 24.9 (3.0) | 3% | 22.6 (3.9) | 8% | 0.3 (2.6x10<sup>-3</sup>) |
| Project Viva | 324 (160, 164) | 26.3 (3.6) | 13% | 24.3 (4.9) | 12% | 0.3 (1.2x10<sup>-6</sup>) |
| RHEA | 94 (45, 49) | 27.2 (4.0) | 20% | 25.1 (5.5) | 16% | 0.3 (1.1x10<sup>-2</sup>) |
| **Total or mean\*** | **4,894 (2,337, 2,530)** | **26.98 (3.2)** | **10%** | **23.7 (4.1)** | **9%** | **0.2** |

\*In the Total row, average BMI and correlation values were calculated by weighting by the inverse variance for each cohort.

### Quality checks

Quality checks of cohort-specific EWAS results are summarised in Supporting Information File S4. Generally, no major problems were identified, but there were a small number of extreme effect estimates in some cohorts, mainly in the sex-stratified models where the sample sizes were lower. Quality checks of the meta-analysis results (Supporting Information File S5) showed that these outliers had little weighting in the meta-analysis and therefore little impact on the EWAS meta-analysis results. Therefore, these values were not excluded from the meta-analysis.

### Associations between paternal BMI and offspring methylation at birth

Table 3 summarises the results of each EWAS meta-analysis model (full results available on our Open Science Framework site at doi:10.17605/OSF.IO/EBTW7). After FDR correction for multiple testing, we did not identify any CpG sites where there was evidence of an association between paternal BMI and offspring DNA methylation at birth (FDR<0.05). Effect estimates for the model with and without adjustment for maternal BMI were very similar: they correlated highly (Spearman’s r=0.97) and the median percentage difference in effect estimates between models was 0.23% (IQR: 0.1%, 0.6%), suggesting that maternal BMI was not a strong confounder. In a sensitivity analysis, excluding cohorts that defined paternal BMI based on maternal-report did not increase the number of CpGs with FDR-corrected P<0.05.

**Table 3.**
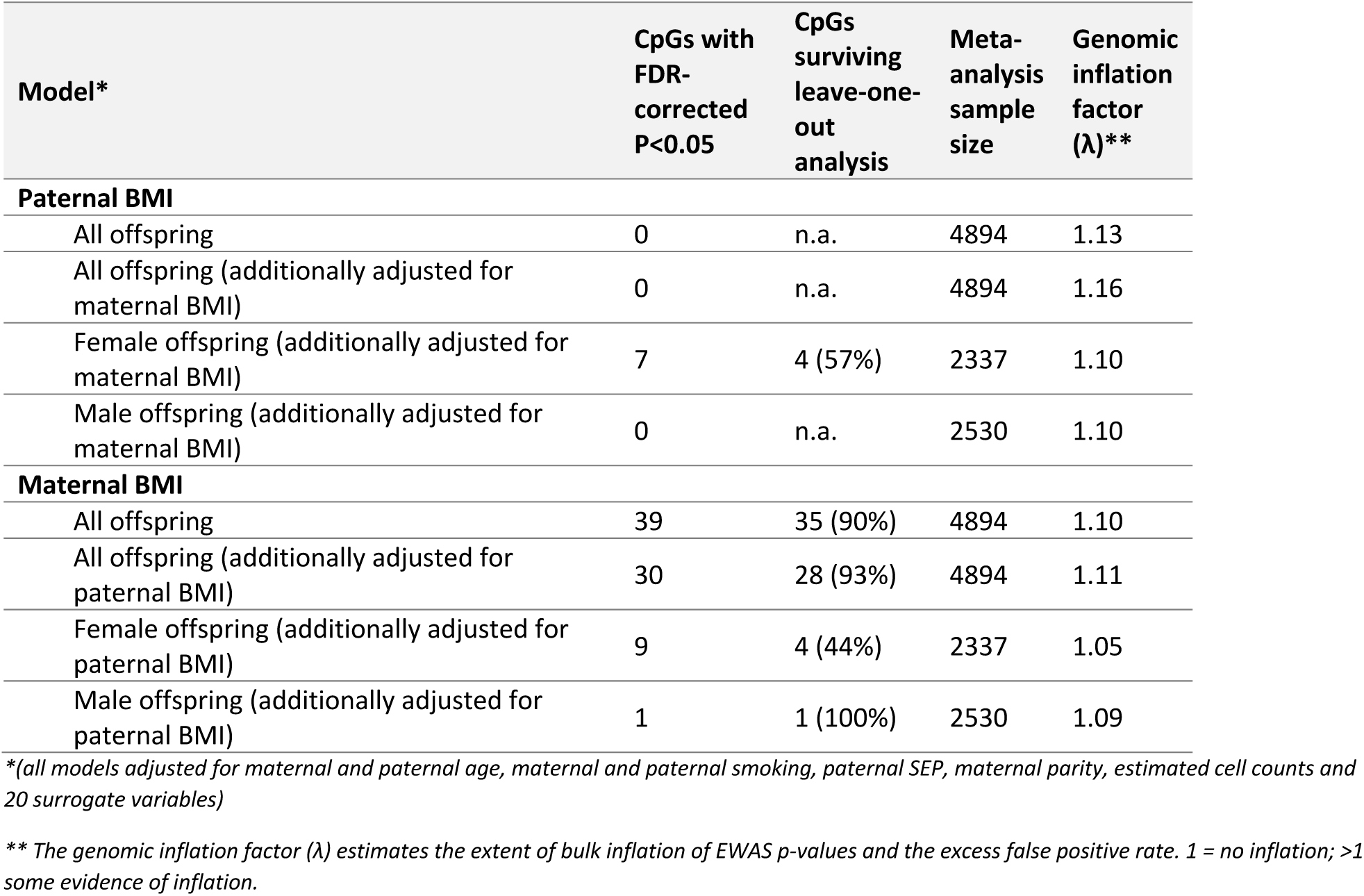
A summary of results of each EWAS meta-analysis model at birth

In a sex-stratified analysis adjusted for maternal BMI, we found some evidence of association between methylation and paternal BMI at seven CpGs in female offspring only (Table 4). Three of these did not survive the leave-one-out analysis (i.e. on omission of one cohort, the effect estimates were in different directions, changed considerably (>20%) and/or had confidence intervals that crossed the null; results in Supporting Information File S6).

**Table 4.**
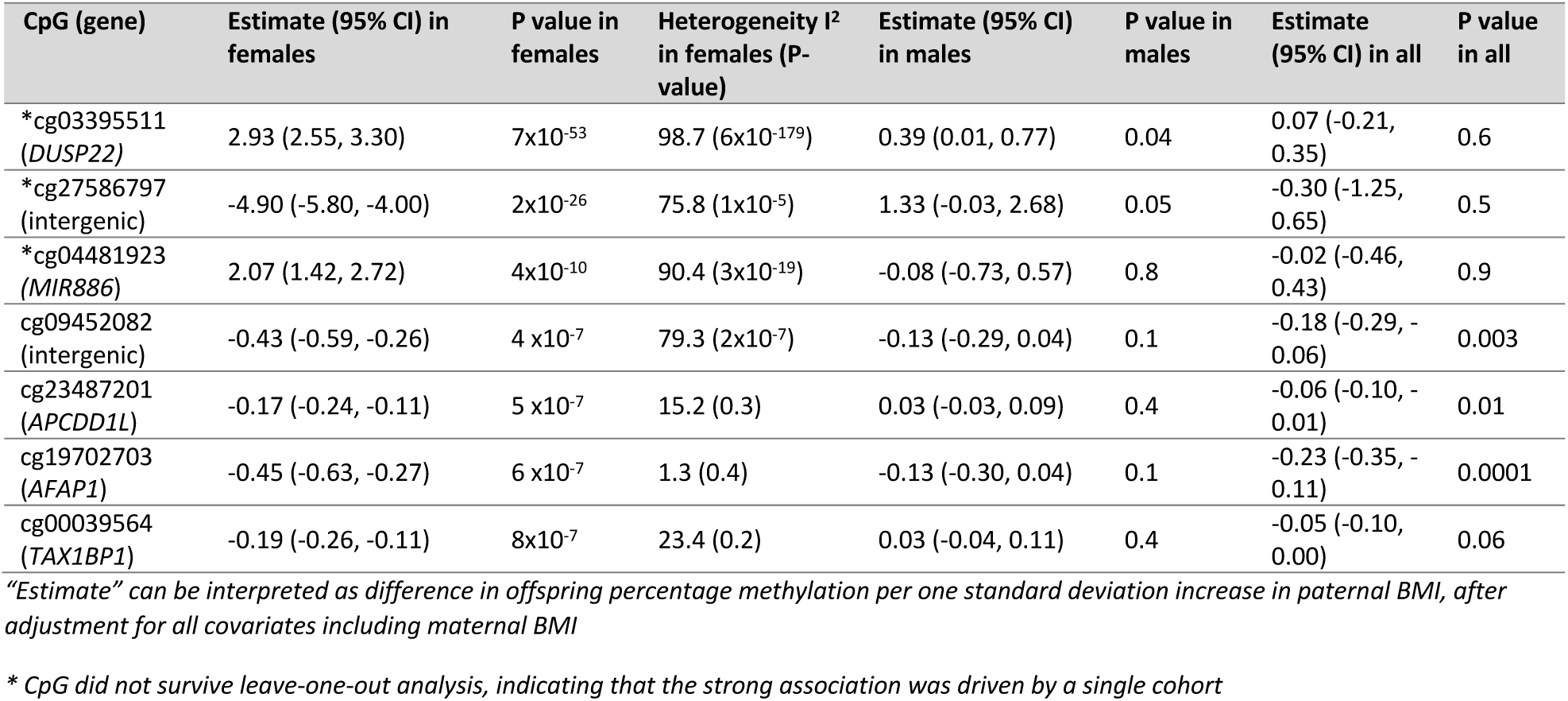
CpGs associated with paternal BMI with FDR-adjusted P<0.05 in female offspring only at birth (estimates are adjusted for maternal BMI)

### Comparison of estimates for paternal and maternal BMI

Maternal BMI was associated with methylation at many more CpG sites than paternal BMI was (Table 3). In the models not stratified by sex, there were 39 CpGs associated with maternal BMI before adjustment for paternal BMI and 30 after (FDR-adjusted P<0.05), 26 overlapped. Most associations survived a leave-one-out sensitivity analysis.

At the top CpGs associated with paternal BMI at a relaxed (but arbitrary) P-value threshold of P<1×10^−5^, the estimated paternal effect was greater (further from the null) than the estimated maternal effect, even after adjustment for the other parents’ BMI (Figure 1). There was also strong evidence of heterogeneity between the maternal and paternal mutually adjusted estimates (all heterogeneity FDR-adjusted P-values<0.05; I^2^ ranging 86.6 to 96.4). However, apart from at the most robustly paternal BMI-associated CpG sites, this pattern (of greater paternal than maternal effect estimates) was not observed. In fact, throughout the genome, around half (49.6%) of CpGs had larger absolute effect estimates for paternal BMI and the other half (50.2%) had larger absolute effect estimates for maternal BMI. Figure 2 shows that maternal effect estimates (before adjustment for paternal BMI) were similar in size to paternal effect estimates, and the distribution of effect estimates across the genome was similar regardless of parent. Results were very similar after mutual adjustment for the other parent’s BMI (Supporting Information File S7).

**Figure 1.**
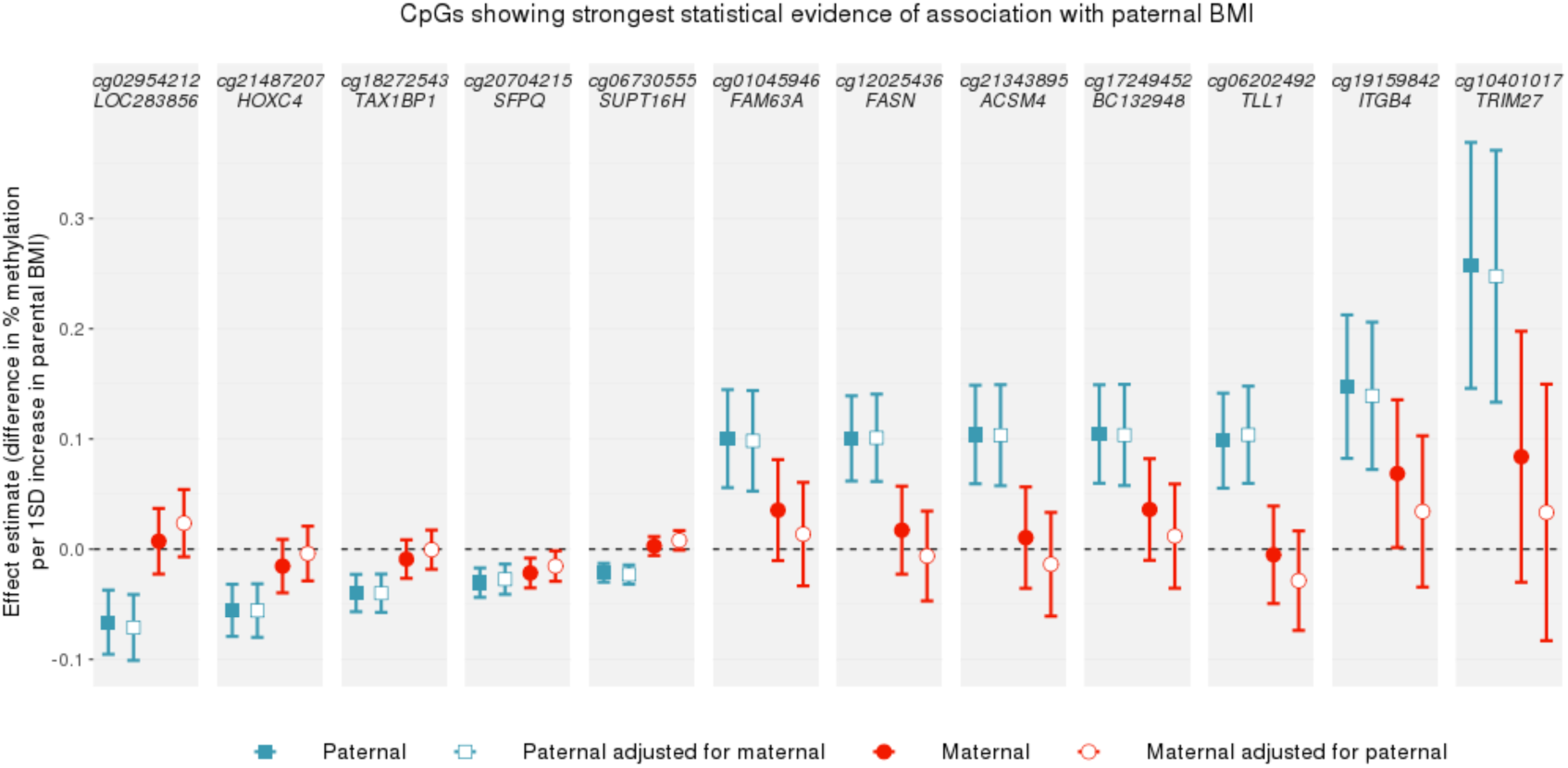
A comparison of paternal and maternal BMI effect estimates at CpGs with P<1*10^−5^ in the paternal BMI EWAS meta-analysis at birth. CpGs were selected if they were associated with paternal BMI with a P-value <1×10^−5^ in the model that was not adjusted for maternal BMI. Points show EWAS meta-analysis effect estimates, bars show 95% confidence intervals. Confidence intervals are not adjusted for multiple testing.

**Figure 2.**
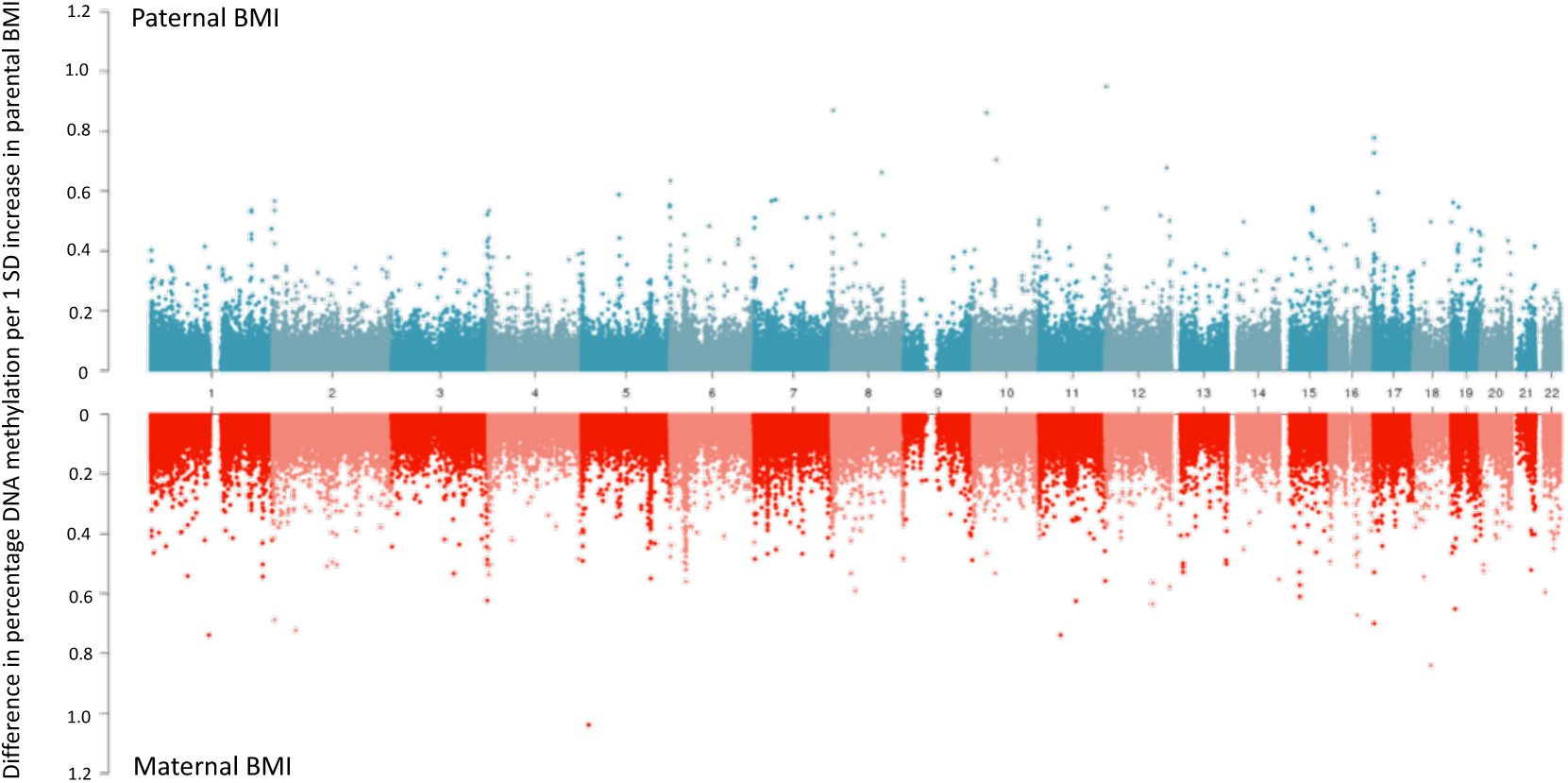
A comparison of paternal and maternal BMI EWAS effect estimates across the genome. Absolute effect estimates (y-axis) are plotted against genomic location (x-axis; numbers indicate chromosome number). Paternal BMI EWAS meta-analysis results are plotted on the top, with maternal EWAS meta-analysis results on a mirrored axis below. Models were not mutually adjusted for the other parent’s BMI (for a comparison of the mutually adjusted results, see Supporting Information File S7).

In a previous PACE consortium study (26), we identified 86 cord blood CpGs associated with maternal BMI in an EWAS meta-analysis across 19 cohorts (nine of which also contributed results to the current study). Of the 86 CpGs identified in that previous study (which had higher statistical power to detect associations), 64 were available in the current study after probe filtering. To explore the extent to which maternal BMI might be driving any association between paternal BMI and offspring methylation in the current study, we assessed enrichment of our paternal BMI EWAS meta-analysis results for these 64 maternal BMI-associated CpGs. We found little evidence of enrichment (Kolmogorov-Smirnov P-value for inflation of EWAS P-values = 0.54 in EWAS unadjusted for maternal BMI; 0.61 in EWAS adjusted for maternal BMI), suggesting that any relationship between paternal BMI and offspring methylation was unlikely to be driven by confounding by maternal BMI, even before adjustment for maternal BMI. Conversely, the maternal BMI EWAS meta-analysis results in the current study were highly enriched for previously-identified maternal BMI-associated CpGs (Kolmogorov-Smirnov P for inflation = 2.2*10^−16^ in EWAS adjusted and unadjusted for paternal BMI). This finding was as expected, given that the main exposure was the same and the samples were overlapping, but it highlights the ability of this analysis to detect strong associations with maternal BMI if they exist.

### EWAS meta-analysis at childhood

#### Cohort summaries

Six cohorts were included in the EWAS meta-analyses at childhood. Table 5 summarises key characteristics of these cohorts. Around 48% of the children were female. In all cohorts, paternal BMI had a higher mean and a lower standard deviation compared to maternal BMI.

**Table 5.** A summary of sex and age of the child and parental BMI for each cohort in the childhood meta-analysis.

| Study | N (N female, N male) | Mean age of children in years (SD) | Mean paternal BMI in kg/m<sup>2</sup> (SD) | Percentage fathers with BMI $\geq 30$ (i.e. obese) | Mean maternal BMI in kg/m<sup>2</sup> (SD) | Percentage mothers with BMI $\geq 30$ (i.e. obese) | Spearman's correlation between paternal and maternal BMI (P-value) |
| --- | --- | --- | --- | --- | --- | --- | --- |
| ALSPAC | 570 (280, 290) | 7.5 (0.2) | 25.0 (3.1) | 7% | 22.6 (3.4) | 4% | 0.2 ($5.1 \times 10^{-6}$) |
| CHAMACOS | 108 (56, 52) | 9.2 (0.3) | 27.6 (3.4) | 27% | 27.0 (4.4) | 22% | 0.2 ($2.9 \times 10^{-2}$) |
| Generation R | 335 (174, 161) | 6.0 (0.4) | 25.0 (3.3) | 6% | 24.1 (3.9) | 6% | 0.2 ($1.5 \times 10^{-5}$) |
| HELIX | 516 (231, 285) | 8.4 (1.7) | 26.6 (3.7) | 18% | 23.9 (4.4) | 9% | 0.3 ($1.1 \times 10^{-10}$) |
| INMA | 177 (87, 90) | 4.4 (0.2) | 26.0 (3.6) | 15% | 24.6 (5.1) | 13% | 0.3 ($9.6 \times 10^{-5}$) |
| Project Viva | 276 (132, 144) | 7.8 (0.7) | 26.4 (3.7) | 15% | 24.5 (4.7) | 13% | 0.4 ($4.2 \times 10^{-13}$) |
| **Total** | **1982 (960, 1022)** | 6.9 (0.3) | **25.7 (3.4)** | **12%** | **24.3 (5.2)** | **9%** | **0.3** |

\* In the Total row, average BMI, age and correlation values were calculated by weighting by the inverse variance for each cohort.

### Quality checks

Quality checks of cohort-specific EWAS results are summarised in Supporting Information File S8. Meta-analysis quality checks are summarised in Supporting Information File S9. We did not exclude any data following these checks.

### Associations between paternal BMI and offspring methylation in childhood

Table 6 summarises the results of each EWAS meta-analysis model (full results available on our Open Science Framework site at doi:10.17605/OSF.IO/EBTW7). There was one CpG where we found evidence (FDR-adjusted P<0.05) of an association with paternal BMI (cg2720130 at *GIP* on chromosome 17; ß 0.4%, 95% CI 0.2% to 0.5%, P 8.8*10^−8^), and one (different) CpG with evidence of an association with maternal BMI (cg07099084 in an intergenic region on chromosome 1, ß −0.05%, 95% CI −0.03% to −0.06%, P 1.1*10^−7^). However, neither survived mutual adjustment for the other parent’s BMI, neither was associated with parental BMI in the birth analysis, and one (cg07099084) did not survive a leave-one-out analysis (Supporting Information File S10). Excluding HELIX from the full meta-analysis did not change the number of associations with FDR-adjusted P<0.05. A meta-regression also showed little evidence that mean age of the children at methylation measurement was associated with EWAS meta-analysis effect estimates (Supporting Information File S10).

**Table 6.** A summary of results of each EWAS meta-analysis model at childhood

| Model* | CpGs with FDR-corrected $P < 0.05$ | CpGs surviving leave-one-out analysis | Meta-analysis sample size | Genomic inflation factor ($\lambda$)** |
| --- | --- | --- | --- | --- |
| **Paternal BMI** |  |  |  |  |
| All offspring | 1 | 1 (100%) | 1982 | 1.03 |
| All offspring (additionally adjusted for maternal BMI) | 0 | n.a. | 1982 | 1.00 |
| Female offspring (additionally adjusted for maternal BMI) | 0 | n.a. | 960 | 1.00 |
| Male offspring (additionally adjusted for maternal BMI) | 0 | n.a. | 1022 | 1.05 |
| **Maternal BMI** |  |  |  |  |
| All offspring | 1 | 0 (0%) | 1982 | 1.11 |
| All offspring (additionally adjusted for paternal BMI) | 0 | n.a. | 1982 | 1.08 |
| Female offspring (additionally adjusted for paternal BMI) | 0 | n.a. | 960 | 1.06 |
| Male offspring (additionally adjusted for paternal BMI) | 0 | n.a. | 1022 | 1.07 |

\*(all models adjusted for maternal and paternal age, maternal and paternal smoking, paternal SEP, maternal parity, estimated cell counts and 20 surrogate variables)
\*\* The genomic inflation factor ( $\lambda$ ) estimates the extent of bulk inflation of EWAS p-values and the excess false positive rate. 1 = no inflation; $>1$ some evidence of inflation.

### Analysis of cell proportions

Paternal BMI was not associated with the proportion of any cell type in offspring blood (Supporting Information File S11), except for a very small difference in the estimated proportion of nucleated red blood cells in offspring cord blood (nRBCs; 0.001 greater proportion of nRBCs per 1SD increase in paternal BMI; 95% CI 0.0004, 0.0017; P=0.001) and an even smaller difference in the proportion of CD4 T-cells in childhood peripheral blood (0.0007 lower proportion of CD4 T-cells per 1 SD increase in paternal BMI; 95% CI −0.0013, −0.0002; P=0.007), which appeared to be largely driven by the HELIX dataset and did not survive a sensitivity analysis excluding HELIX.

### Systematic literature review

Figure 3 summarises the workflow and Table 7 outlines the seven included studies resulting from a systematic literature review of human studies of paternal adiposity and offspring or gamete methylation. There were five studies of imprinted regions and two untargeted array-based studies. Four studies investigated DNA methylation in offspring cord blood, two in paternal sperm and one in both.

**Table 7.** Summary of identified studies of paternal adiposity and sperm or offspring DNA methylation. In all studies of offspring methylation, the estimated effect of paternal BMI/obesity was adjusted for maternal BMI/obesity.

| Study | Exposure | Outcome | Sample size | Studied regions | Key findings |
| --- | --- | --- | --- | --- | --- |
| Noor et al. 2019 (20) | Paternal BMI | Offspring DNA methylation at birth (cord blood), age 3 and age 7 (peripheral blood) | 429 | Untargeted (Illumina 450k array) | Paternal BMI was associated with cord blood DNA methylation at 9 CpGs with an FDR-adjusted P-value <0.05. Three of these persisted at age 3 and one of those also persisted at age 7. When stratified by maternal BMI, no CpGs were associated with paternal BMI in the subset with maternal BMI <25, but 18 CpGs were identified in the subset with maternal BMI ≥25. None persisted at later time points. |
| Potabattula et al. 2019 (19) | Paternal BMI | Sperm DNA methylation and cord blood DNA methylation | 294 (sperm) and 113 (cord blood) | Imprinted genes: *MEST/PEG1*, *SNRPN/PEG4*, *NNAT/PEG5*, *SGCE/PEG10*, *H19-IG*, *IGF2*, and *MEG3-IG*, and one obesity related gene: *HIF3A* | Paternal BMI was positively associated with *MEG3* methylation in sperm and in male (but not female) offspring. There were some other small magnitude associations in sex-stratified analyses. |
| Potabattula et al. 2018(17) | Paternal BMI | Cord blood DNA methylation | 46 | Six imprinted genes: *H19*, *IGF2*, *MEST*, *PEG3*, *MEG3*, *NNAT* | Paternal BMI was positively associated with methylation of the paternal *MEST* allele. |
| Soubry et al. 2016(16) | Paternal overweight or obesity | Sperm DNA methylation | 67 (23 overweight or obese) | 12 imprinted genes: *MEG3*, *MEG3-IG*, *IGF2*, *H19*, *GRB10*, *NDN*, *NNAT*, *PLAGL1*, *SGCE/PEG10*, *SNRPN*, *PEG1/MEST*, *PEG3* | Paternal overweight/obesity was associated with lower sperm methylation at *MEG3*, *NDN*, *SNRPN* and *SGCE/PEG10* and higher methylation at *MEG3-IG*, *H19*, *IGF2*, compared to controls. There was little evidence of association at *GRB10*, *NNAT*, *PLAGL1*, *PEG1/MEST* and *PEG3*. |
| Donkin et al. 2015(18) | Paternal obesity | Sperm DNA methylation | 23 (10 obese) | Untargeted (RRBS) | There were 9,081 unique genes differentially methylated in the sperm of lean compared to obese men with an FDR-adjusted P<0.1 (7,059 with FDR-adjusted P<0.05). |
| Soubry et al. 2015(15) | Paternal obesity and paternal BMI | Cord blood DNA methylation | 63 (16 obese) | Seven imprinted genes: *MEG3*, *MEST*, *NNAT*, *PEG3*, *PLAGL1*, *SGCE*, *PEG10* | Paternal obesity was associated with lower offspring methylation at *MEST*, *NNAT* and *PEG3*. Paternal BMI was positively correlated with methylation at *SGCE/PEG10*. |
| Soubry et al. 2013(14) | Paternal obesity | Cord blood DNA methylation | 70 (16 obese) | Two imprinted genes: *H19*, *IGF2* | Paternal obesity was associated with lower methylation at *IGF2* compared to controls, but there was no difference at *H19*. |

RRBS: reduced representation bisulfite sequencing

**Figure 3.**
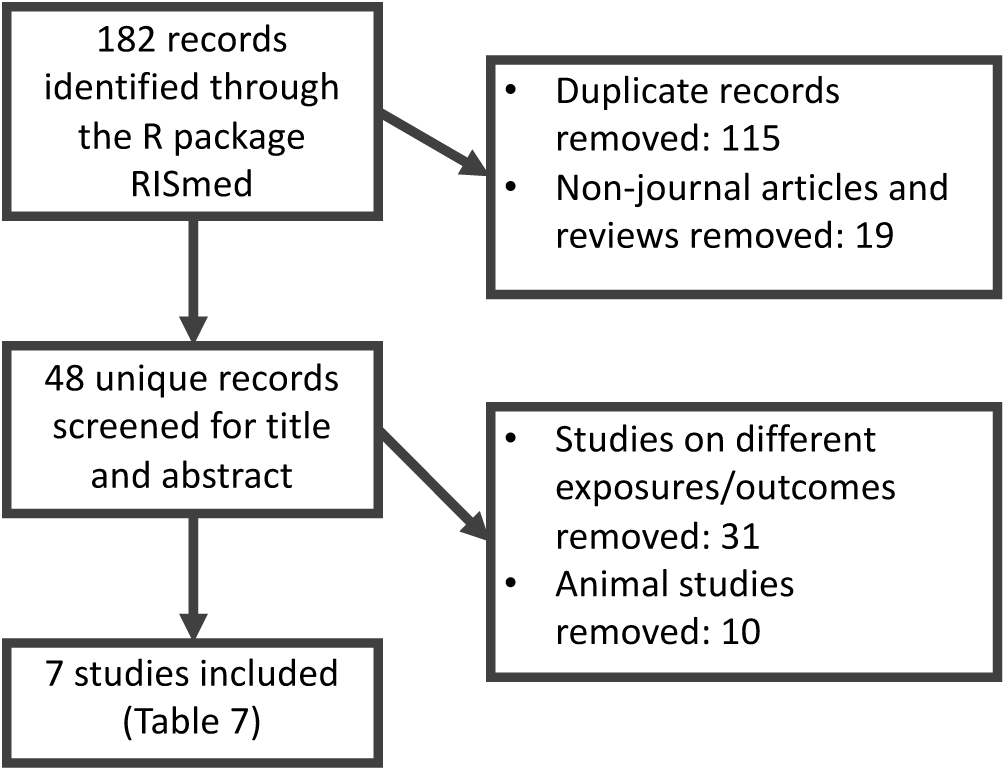
The systematic review process used to identify human studies of paternal adiposity and offspring or germ cell methylation

### Comparison to the literature

#### Imprinted regions identified in studies by Soubry et al. and Potabattula et al

At paternally-imprinted regions identified in the literature review, EWAS meta-analysis effect estimates for the association between paternal BMI and cord blood methylation (adjusted for maternal BMI) were small with no clear trend in direction (Figure 4). Findings were similar for the EWAS model unadjusted for maternal BMI (Supporting Information File S12).

**Figure 4.**
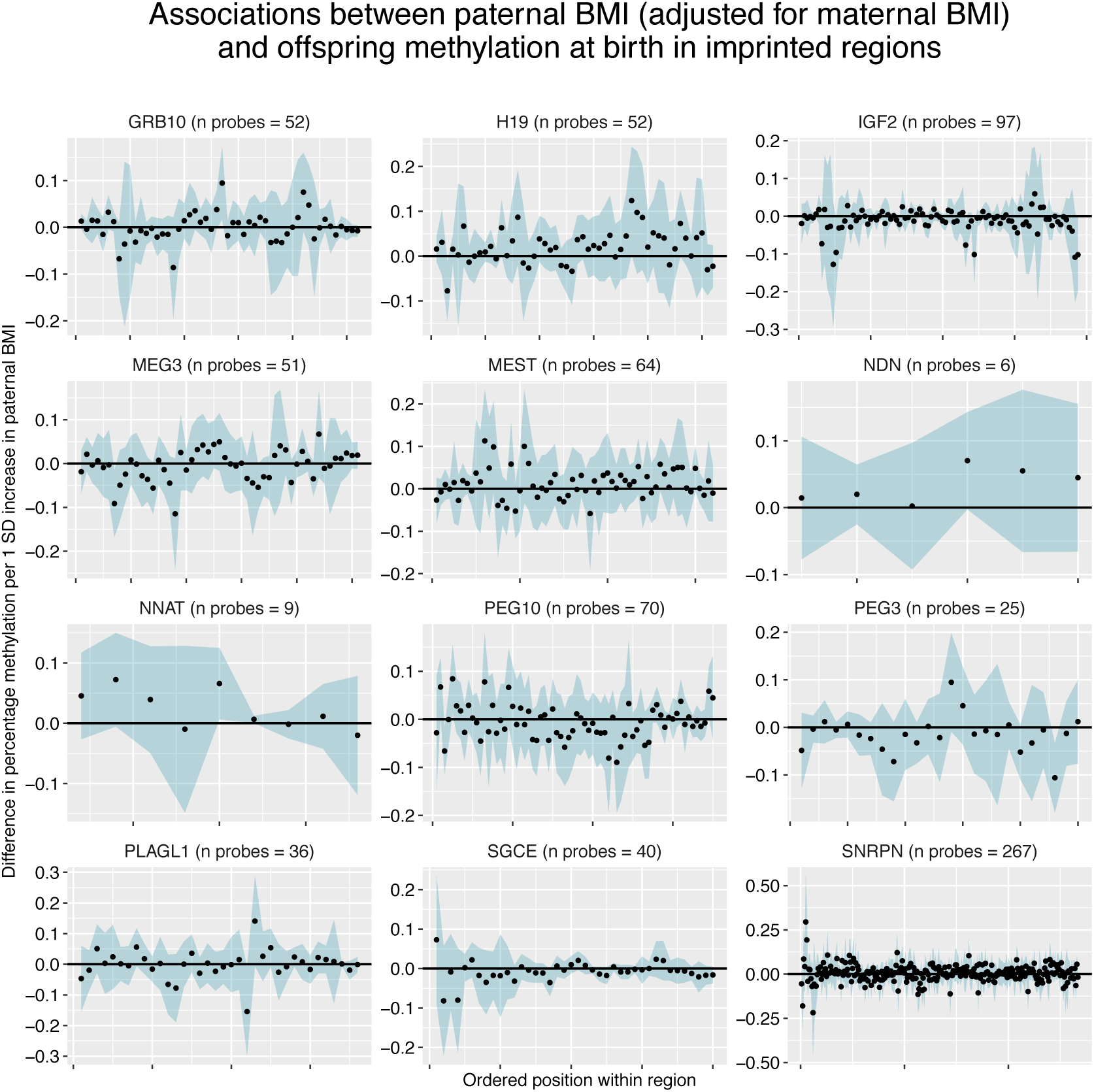
Paternal BMI effect estimates at CpGs within imprinted regions. Each panel shows a different imprinted gene, with CpGs arranged in order on the x-axis. The blue ribbon shows the 95% confidence intervals. All results are adjusted for maternal BMI.

### Regions identified by Donkin et al

Donkin et al. (18) reported 9,081 genes differentially methylated between sperm samples from lean and obese men (with FDR-adjusted P<0.1). Of these, we could only map to 511 CpGs at the same genomic positions in our cord blood EWAS meta-analysis results, because Donkin et al. measured methylation using reduced representation bisulfite sequencing (RRBS) with a higher coverage of the genome than the 450k array. In our paternal BMI EWAS meta-analysis at birth (adjusted for maternal BMI), we found the same direction of effect at only roughly half of these genes (252/511), and only 17 of these had a P-value<0.05, with none surviving FDR correction for multiple testing at either 511 or 252 sites. QQ plots and a Kolmogorov-Smirnov test suggested that our EWAS P-values at these 252 sites were not smaller than would be expected by chance (Supporting Information File S12; Kolmogorov-Smirnov P = 0.27). Findings were similar when using the EWAS meta-analysis P-values from the paternal BMI model that was not adjusted for maternal BMI (Supporting Information File S12).

### CpGs identified by Noor et al

In Project Viva (a cohort that also contributed results to our meta-analysis), Noor et al. identified nine CpGs where cord blood methylation was associated with periconceptional paternal BMI after adjustment for maternal BMI (20). In our results at birth, we found the same direction of estimated effect at 7/9 CpGs. Only one of these had a P-value<0.05, but this association also survived FDR correction for multiple testing at nine sites (cg04763273 mapping to *TFAP2C*; −0.41% difference in methylation per 1SD increase in paternal BMI; 95% CI −0.67 to −0.15; P=0.002). According to our leave-one-out analysis criteria, this CpG “survived” after omission of Project Viva, but it did not survive after omission of Generation R. A Kolmogorov-Smirnov test suggested that our EWAS P-values at the nine CpGs identified by Noor et al. were not smaller than would be expected by chance (Kolmogorov-Smirnov P=0.21). Noor et al. also identified 18 CpGs where cord blood methylation was associated with paternal BMI when the analysis was restricted to a subset of offspring of mothers with a BMI>25. In our (unstratified) results at birth we found the same direction of estimated effect at only nine of these 18 CpGs, only one had a P-value<0.05 (cg04763273) and a Kolmogorov-Smirnov test suggested our EWAS P-values were not smaller than would be expected by chance (Kolmogorov-Smirnov P=0.54).

## Discussion

### Summary and interpretation of findings

In coordinated EWAS meta-analysis using a total of 19 datasets, at a genome-wide level of significance (FDR-adjusted P<0.05) we found little evidence of association between prenatal paternal BMI and offspring blood DNA methylation at birth or in childhood. In sex-stratified analyses, we found robust associations between paternal BMI and cord blood methylation at just four CpGs in female offspring only. There was little evidence of residual confounding by maternal BMI, and paternal BMI was not strongly associated with estimated cell heterogeneity at either timepoint. At the top CpGs most strongly associated with paternal BMI, estimates of the effect of paternal BMI on methylation were larger than estimates of the effect of maternal BMI, suggesting some evidence of paternal-specific effects at these top CpGs. However, more associations with maternal BMI than paternal BMI surpassed our P-value threshold (FDR-adjusted P<0.05), and across the whole genome estimates of the effect of paternal BMI were similar in magnitude to those of maternal BMI.

The lack of evidence for large numbers of associations with low P-values is partly influenced by limited power and we cannot discount an impact of paternal BMI on offspring DNA methylation without larger numbers and/or studies in populations with greater variability in paternal BMI. Furthermore, we cannot rule out an impact on DNA methylation measured in different tissues and/or at regions not covered by the 450k array.

### Comparison to the literature

Our null findings are in contrast to some previous findings. In a systematic review, we identified seven studies that have previously reported on associations between paternal BMI and sperm or offspring methylation in humans. Five of these (three from one study and group(14–16) and two from another (17,19)) were candidate gene studies that found associations between paternal obesity or higher BMI and sperm or offspring cord blood methylation at imprinted regions (all after adjustment for maternal BMI or obesity). There is a strong biological rationale to studying imprinted regions in the context of paternal exposures, because methylation marks at imprinted regions appear to survive the wave of demethylation that occurs following fertilization, and therefore have the potential to pass on epigenetic information from the gametes to the offspring(10,59). However, where studies of imprinted genes have considered the same loci, the direction of estimated effects have been discordant between studies. For example, Potabattula et al.(17) found a positive correlation between paternal BMI and cord blood methylation at *MEST*, but Soubry et al.(14) found that cord blood of offspring of obese fathers was lower than that of controls, indicating a negative correlation with BMI. There are also paradoxical findings at *SGCE/PEG10 and IGF2* between studies of sperm(16) and cord blood(14), but if methylation differences in sperm are transmitted to offspring, we would expect the same direction of estimated effects in these tissues. In our EWAS meta-analysis of paternal BMI, we found little evidence of enrichment for imprinted genes. Effect estimates at individual CpGs within imprinted regions were small and the direction of effect varied within most genes (Figure 4). Therefore, we found little evidence to support the findings of these candidate gene studies, despite having a much larger sample size and statistical power. However, it should be noted that the Illumina BeadChip array does not provide full coverage of imprinted regions, so further, more detailed analysis of these regions may be justified.

One small study (total n=23; 10 obese) by Donkin et al.(18), used an untargeted genome-wide approach that identified over 9,000 CpGs throughout the genome differentially methylated in the sperm of obese versus lean men. Only 511 of these CpGs were available in our meta-analysis, but we found little evidence of association at these. The difference in findings of the two studies could reflect a number of factors, including differences in the studied tissue (sperm vs offspring blood), technology (RRBS versus 450k), definition of phenotype (i.e. obese/lean versus BMI over the whole range), or study sample (e.g. all the obese men in Donkin et al. were glucose intolerant).

Finally, in our meta-analysis we replicated the direction of effect at seven out of nine CpGs identified in Noor et al.’s study of periconceptional paternal BMI and cord blood DNA methylation in Project Viva (20). One of these CpGs (cg04763273 near *TFAP2C*) appeared robustly associated with paternal BMI, but we found limited evidence to support their other findings. Our study designs were similar in that we used the same Illumina array, exposure definition, offspring tissue and timepoint, and even had some sample overlap (Project Viva contributed results to our meta-analysis). However, there were some differences in the model and our sample size was over 11 times larger and therefore more robust to identifying false positives.

### Strengths

Analyses were conducted according to a pre-specified, harmonised analysis plan, which is publicly available to aid reproducibility. All the code used to conduct these analyses, and all the resulting EWAS meta-analysis summary statistics, are publicly available on our Open Science Framework site at doi:10.17605/OSF.IO/EBTW7. This means that new studies with relevant data could undertake identical analyses and meta-analyse with our results to produce more precise estimates.

Our study uses a sample size of 4,894, which is >11-fold larger than all the independent studies identified in our systematic review (Table 7), the largest of which has a sample size of 429. It draws together rich data from multiple birth cohorts internationally and this richness allowed us to adjust for important potential confounders. We also attempted to adjust for systematic variation in the DNA methylation data by generating and adjusting for surrogate variables, which is an approach that has been shown to reduce the risk of false positives(44). In a further attempt to ensure the robustness of our results, we conducted sensitivity analyses (leave-one-out at top sites, excluding cohorts with only maternal-reported paternal BMI, excluding HELIX from the genome-wide meta-analysis, and meta-regression of age).

Our rich data enabled us to conduct a series of novel analyses. Firstly, we explored associations between paternal prenatal BMI and offspring DNA methylation at childhood, which allows exploration of the persistence of associations from birth, and/or the effect of paternal BMI postnatally (which we would expect to be correlated with prenatal paternal BMI). Secondly, we explored associations between paternal BMI and cellular heterogeneity, which is an important source of variation in methylation data, but also an interesting phenotype to study in its own right (54). Thirdly, we have previously shown that maternal BMI is associated with offspring methylation, which may reflect a causal intrauterine effect at some CpGs (26). Therefore, to help us tease apart paternal from maternal effects, we compared paternal models adjusted and unadjusted for maternal BMI and we also studied maternal BMI as the main exposure. A particular advantage here was that we included the same samples in both analyses and adjusted for the same covariates, so the sample sizes were the same and the main exposure (paternal or maternal BMI) was the only difference between models. We calculated the correlation between maternal and paternal BMI in each cohort which provided modest support for assortative mating, but by comparing paternal and maternal effect estimates and assessing enrichment of our paternal results for the CpGs we previously identified as associated with maternal BMI (26), we were able to infer that maternal BMI was not an important confounder driving associations between paternal BMI and offspring methylation.

### Limitations

There are a number of limitations that should be considered when interpreting our results. Firstly, the 450k array covers only 1.7% of CpGs on the genome, therefore regions (including imprinted regions) that might be differentially methylated in association with paternal BMI might be missed. Secondly, BMI could be a poor measure of paternal adiposity in our sample, partly due to general issues with the use of BMI as a measure of adiposity(60,61) (perhaps particularly in men) and partly due to measurement error introduced by self- or partner-report. One study showed that lean mass explained more variability in men’s BMI than body fat did, whereas the opposite was seen in women(60). Measurement error might be higher for paternal BMI than maternal BMI because few cohorts had direct measurements and in some instances BMI was reported for the father by the mother, introducing additional measurement error to the questionnaire reported variable (however, we found similar results in our EWAS meta-analysis when we excluded cohorts that used maternal-report to define paternal BMI). More variation in offspring methylation might be captured by another measure such as fat mass percentage or a measure of “lipotoxicity” (61), but these data were not available. Thirdly, offspring blood might not be the most suitable tissue to study and effects might be seen in other tissues such as adipose (given previous evidence of associations between paternal and offspring adiposity), but again, we were limited by data availability. Fourthly, the range of paternal BMI in our study sample could be insufficient to show an effect. As with associations between maternal BMI and offspring adiposity (62), there might be a J- or U-shaped relationship between paternal BMI and offspring outcomes, whereby strong associations are only seen at the extremes of the distribution. We anticipated that we would not have enough power to dichotomise into (extreme) obese and lean groups, therefore we were limited to only studying paternal BMI across the whole range as a continuous variable. Indeed, in our birth analysis, only 10% of fathers across all cohorts were obese (Table 2; Supporting Information File 3), which is the same as a recent estimate of the worldwide prevalence of obesity in men, but much lower than the estimated prevalence of obesity in adults living in Europe (23%) and America (28%)(63), where most of our sample reside. This relatively low occurrence of paternal obesity (and therefore variability in BMI) in our sample may partly explain why we did not find stronger evidence of association with our outcome. Finally, paternal data, including paternal BMI, from birth cohort studies is potentially more at risk of bias than maternal data. Lower prioritisation and greater difficulties recruiting fathers can introduce a higher degree of missing data for fathers compared to mothers. If paternal participation is related to BMI, this could create a selection bias in our sample. Additionally, non-paternity (i.e. partners not being genetically related to their offspring) might also introduce bias if it is related to paternal BMI. Both of these issues could bias paternal estimates towards the null and therefore could be a plausible non-biological reason for our observation of more FDR<0.05 associations between offspring methylation and maternal than paternal BMI.

### Suggestions for further research

Paternal effects on offspring health have been observed, so there is a logical motivation to further investigate the potential underlying molecular mechanisms. As mentioned above, in studies of the effect of paternal adiposity, this work could focus on extreme ends of the BMI distribution and/or other more informative measures of adiposity such as fat mass index, waist-circumference or measures of lipotoxicity (61). There is also huge scope to study the effects of other paternal characteristics, such as health behaviours like smoking and alcohol consumption. Evidence from animal studies suggests that other epigenetic mechanisms such as long non-coding RNAs or tRNAs could be more likely intergenerational carriers of paternal information than DNA methylation and it would be useful to extend these investigations to humans.

## Conclusion

In this large EWAS meta-analysis, we found little evidence of association between paternal pre-pregnancy BMI and offspring DNA methylation in blood, including at imprinted genes. However, this does not rule out the possibility of a paternal epigenetic effect in different tissues or via different mechanisms. More research is warranted to gain a greater understanding of the size and nature of contributions of paternal adiposity to offspring outcomes more broadly.

## Data Availability

All code used to generate our results is available on our Open Science Framework site at doi:10.17605/OSF.IO/EBTW7. We are not able to publicly share individual level data from participating cohorts due to issues with consent and ethics, however all summary statistics generated by meta-analysis will be available at doi:10.17605/OSF.IO/EBTW7 once this pre-print has been accepted for publication in a peer-reviewed journal.

https://osf.io/ebtw7/

## Acknowledgments

Acknowledgments and funding for each cohort and research group are listed in Supporting Information File S1.

## Supporting Information

File S1) Cohort-specific methods, funding and acknowledgements

File S2) Literature review search strategy

File S3) Summary of cohort variables

File S4) Quality checks of cohort-specific EWAS results at birth

File S5) Quality checks of meta-analysis EWAS results at birth

File S6) Results of sensitivity analyses at birth

Files S7) Comparison of paternal and maternal BMI EWAS results after adjustment for maternal BMI

File S8) Quality checks of cohort-specific EWAS results at childhood

File S9) Quality checks of meta-analysis EWAS results at childhood

File S10) Results of sensitivity analyses at childhood

File S11) Results of the meta-analysis of paternal BMI in relation to cellular heterogeneity

File S12) Additional results of the comparison to the literature

